# Trajectories of depressive symptoms across pregnancy and the extended postpartum period and future cardiovascular health

**DOI:** 10.64898/2026.05.26.26353833

**Authors:** Shannon D. Donofry, Megan M. McLaughlin, Emily S. Miller, William Grobman, George Saade, Neil J. Wimmer, Matthew K. Hoffman, Lauren Theilen, Lynn M. Yee, C. Noel Bairey Merz, Caroline E. Rouse, Jessica M. Page, Kelly Zafman, Alexandra Berra, Janet M. Catov

## Abstract

**Background:** Individuals diagnosed with depression during pregnancy are more likely to develop cardiovascular disease (CVD) later in life. However, it remains unclear whether subclinical depressive symptoms or symptom trajectories across time are associated with indicators of cardiovascular health (CVH). Therefore, the present study evaluated the relationship between longitudinal depressive symptom trajectories beginning in pregnancy and future CVH.

**Methods:** This secondary analysis of the multisite prospective nuMoM2b-Heart Health Study and included participants with complete longitudinal data from early pregnancy to 2-7 years post-delivery. Participants self-reported depressive symptoms using the Edinburgh Postnatal Depression Scale (EPDS) at 6-13 weeks gestation (early pregnancy), 22-29 weeks gestation (mid- to late-pregnancy), and 2-7 years post-delivery. Latent class mixture modeling was conducted to identify longitudinal patterns of depressive symptoms across early pregnancy, mid-late pregnancy, and extended postpartum follow-up. Structural equation modeling was used to test whether EPDS trajectories were associated with latent CVH, adjusted for length of follow-up interval, pre-pregnancy BMI, gravidity, adverse pregnancy outcomes, smoking history, age, education, income, and use of psychiatric medications.

**Results:** A total of 3,934 participants (mean (*M*) ± standard deviation (*SD*) age=27.6±5.6 years) met inclusion criteria with a mean follow-up interval of 3.2±0.9 years. A 4-class model, which provided the best fit to the EPDS data (mean posterior probability across classes=0.81), produced the following trajectories: (1) stable low (n=2412; 61.1%), (2) increasing severity (n=848; 21.5%), (3) decreasing severity (n=476; 12.1%), and (4) stable high (n=212; 5.4%). Compared to the stable low group, all groups exhibited significantly lower CVH (stable high: β=0.06, *p*<0.01; decreasing severity: β=0.05, *p*=0.02; increasing severity: β=0.08 *p*<0.01). Pairwise comparisons among the three elevated-symptom groups revealed no significant differences in latent CVH (all *p*s >0.24).

**Discussion:** The longitudinal course of depressive symptoms from pregnancy to 2-7 years post-delivery varied across individuals. Compared to those with consistently low depressive symptoms, individuals with higher severity symptoms at any point all exhibited lower CVH, regardless of the specific trajectory of symptoms. These findings support a life-course perspective in which depressive symptom patterns may represent an early indicator of cardiometabolic vulnerability.

**Clinical Perspective:** **What is New?**

- We examined the longitudinal relationship between differing trajectories of depressive symptoms during pregnancy and throughout the extended postpartum period and future cardiovascular health.
- We found that individuals with stably elevated, decreasing severity and increasing severity depressive symptoms from pregnancy to 2-7 years post-delivery exhibited lower cardiovascular health 2-7 years post-delivery relative to individuals with stably low depressive symptoms.

**What are the Clinical Implications?**

- The longitudinal course depressive symptoms from pregnancy through the extended postpartum period varies across individuals, and individuals with consistently elevated, elevated with decreasing severity, and low with increasing severity symptoms exhibit poorer cardiovascular health compared to those with consistently low severity symptoms. This suggests that any elevation of depressive symptoms, regardless of whether they exceed clinical thresholds indicative of a depressive episode, may relate to cardiovascular risk.
- Routine assessment of depressive symptoms across pregnancy and in the years following delivery may facilitate early detection of individuals who may be at risk of CVD in the future.

## Introduction

Pregnancy represents a period of heightened vulnerability for the onset or exacerbation of depressive symptoms, with approximately 10-20% of women experiencing clinically significant depression during the perinatal period^1–4^. Antenatal depression is one of the strongest predictors of postpartum depression^5,6^, and individuals who experience depression during the perinatal period are at elevated risk of future depressive episodes^7^. Further, antenatal depression is associated with numerous adverse pregnancy outcomes (APOs), including increased risk of preterm birth, low birth weight, pre-eclampsia^8–12^, and these pregnancy complications are now recognized as sex-specific risk enhancers for cardiovascular disease (CVD)^13–15^. Yet, despite the well-documented associations between depression and CVD in non-pregnant populations^16–20^, relatively few studies have directly examined whether depressive symptoms during pregnancy and the extended postpartum period related to future cardiovascular health (CVH). The limited available evidence, drawn primarily from large population-based registries, suggests that women diagnosed with antenatal depression demonstrate increased rates of CVD events in subsequent years^21–23^. For example, two large longitudinal studies demonstrated that diagnosed antenatal depression was associated with increased risk of ischemic heart disease, cardiac arrest, and cardiomyopathy within the first two years postpartum^21^ and more than double the risk of incident CVD two decades later^22^.

Nevertheless, prior studies examining the link between depression in pregnancy and future CVD risk have been constrained by several important limitations. First, existing research has focused predominantly on clinically diagnosed depression, leaving unclear whether subclinical depressive symptoms, which affect a substantially larger proportion of pregnant individuals^24,25^, are also associated with CVD-relevant outcomes. Second, prior investigations have largely assessed antenatal depression as a static exposure, failing to account for the substantial heterogeneity in symptom trajectories during and after pregnancy. Some individuals experience persistently elevated symptoms throughout pregnancy and postpartum, while others demonstrate worsening or improving symptom patterns over time^10,26–28^. Recent evidence from non-pregnant populations suggests that CVD risk varies based on patterns of depressive symptom trajectories over time^29,30^. As such, distinct trajectories during pregnancy and the extended postpartum period may similarly confer differential risk for future cardiovascular outcomes.

The present study aims to address these knowledge gaps by examining associations between trajectories of perinatal depressive symptoms and future cardiovascular outcomes, using data from the Nulliparous Pregnancy Outcomes Study: Monitoring Mothers-to-Be (nuMoM2b) and nuMom2b-Heart Health Study (nuMoM2b-HHS). We hypothesized that women with persistently elevated or worsening depressive symptoms across pregnancy and the extended postpartum period would demonstrate worse CVH later in life.

## Methods

### Participants and study procedures

This was a secondary analysis of nuMoM2b, a large multisite prospective pregnancy study conducted at eight US medical centers, and nuMoM2b-HHS, a follow-up cohort study of nuMoM2b participants conducted 2-7 years after delivery. Both studies were approved by all local institutional review boards, and all participants provided written informed consent prior to the onset of any study procedures. Additional details about participants and methodological protocols can be found in prior publications.^31,32^

Eligible nuMoM2b participants completed assessments at multiple timepoints across pregnancy: 6-13 weeks’ gestation (hereafter referred to as V1), 16-21 weeks’ gestation (early-mid second trimester; V2), 22-29 weeks’ gestation (late second-early third trimester; V3), and at delivery (V4). After delivery, information about birth outcomes and pregnancy complications was abstracted from medical records. Data collection for nuMoM2b occurred from 2010 to 2013.

NuMoM2b participants were eligible for the in-person nuMoM2b-HHS visit if they were at least 18 years of age, had delivery data on the index pregnancy, were at least six months postpartum from any subsequent pregnancy, and were not currently pregnant. Of the original 10,038 nuMoM2b participants, a total of 7,003 were successfully contacted for follow-up, and 4,508 completed an in-person assessment 2-7 years after delivery (hereafter referred to as V5). At the in-person visit, trained personnel collected clinical measurements and blood samples, and participants self-reported information about their medical and psychosocial history. Sociodemographic characteristics of individuals who did and did not complete the in-person visit were similar.^33^

The present study included 3,947 participants (M_age_=27.6±5.6 years) with complete longitudinal data for variables of interest from the first trimester of pregnancy to 2-7 years post-delivery (V1, V3, and V5). Figure 1 depicts a flow diagram for participant inclusion in the analysis.

**Figure 1.**
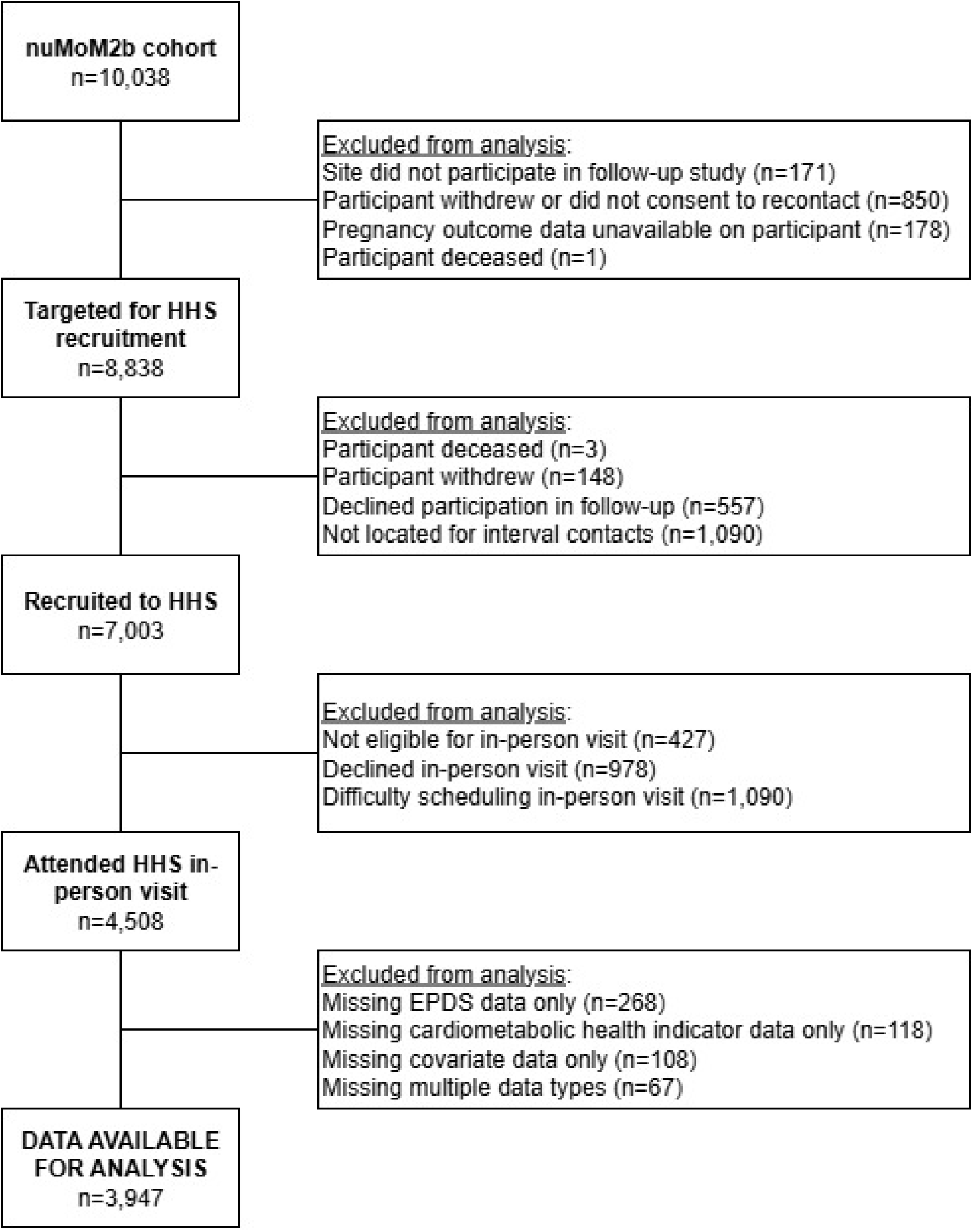
Flow diagram for participation in nuMoM2b and nuMoM2b-HHS, as well as for inclusion in the current analysis. *Note.* nuMoM2b = Nulliparous Pregnancy Outcomes Study: Monitoring Mothers-to-Be; HHS = Heart Health Study; EPDS = Edinburgh Postnatal Depression Scale.

### Measures

#### Sociodemographic and health characteristics

Participants self-reported sociodemographic characteristics during V1. These included age, education, household income, and racial and ethnic identity reported using standard options. Lifetime tobacco use history was self-reported. Measured height at V1 and self-reported pre-pregnancy weight were used to estimate pre-pregnancy BMI (kg/m^2^). Finally, participants reported the number of times they have been pregnant, including the index pregnancy.

#### Depressive symptoms

The presence and severity of depressive symptoms were measured using the 10-item Edinburgh Postnatal Depression Scale (EPDS)^34^, administered at three timepoints: V1, V3, and V5. The EPDS is a widely used instrument validated in pregnancy and the postpartum period^35,36^, as well as among non-pregnant women^37^. Respondents rate the severity of depressive symptoms experienced in the past week on a Likert scale ranging from 0 to 3. Scores are summed up to form a total score, with higher scores reflecting more severe symptomatology. A score of ≥13 is indicative of risk for a major depressive episode.^34,35^ The EPDS has demonstrated adequate internal consistency in prior research (Cronbach’s α = 0.77)^34^.

#### Cardiovascular Health Measures

The present study focused on CVH outcomes measured during the in-person nuMoM2b-HHS visit. Informed by prior work by Marsland and colleagues^38^, the outcomes of interest for this analysis included systolic and diastolic blood pressure (mmHg), high-density lipoprotein (HDL) cholesterol (mg/dL), triglycerides (mg/dL), insulin (uIU/mL), glucose (mg/dL), BMI (kg/m2), and waist circumference (cm). Trained personnel obtained three resting blood pressure measurements using a calibrated automated device (OMRON HEM-907XL, Omron Healthcare Incorporated, Lake Forest, Illinois) while the participant was seated. The average of the last 2 systolic and diastolic pressures were used to index blood pressure. Height was measured using a stadiometer and weight was obtained using a regularly calibrated balance-beam scale, and these data were used to calculate BMI. Waist circumference was measured twice over the iliac crest to the nearest 0.1 cm, using a non-stretch measuring tape. Blood samples were taken via venipuncture and lipids, glucose, and insulin were measured via serum stored at −80°C and batched processed at a core laboratory (The Lundquist Institute) using enzymatic assays^33^.

#### Adverse pregnancy outcomes

Data on APOs were abstracted by study staff from participant medical records, and included gestational diabetes, hypertensive disorders of pregnancy, preterm birth, small-for-gestational-age birth weight, and placental abruption. Gestational diabetes was diagnosed using clinical criteria. Hypertensive disorders of pregnancy included gestational hypertension, pre-eclampsia, eclampsia, and HELLP (hemolysis, elevated liver enzymes, low platelets) syndrome^39^. Preterm birth was defined as any delivery occurring before 37 weeks gestation based on the best obstetric estimate^40^ and included both spontaneous and medically indicated pre-term birth. Small-for-gestational-age birth was defined as below the 10^th^ percentile according to norms obtained from nationally representative birth data^41^. Placental abruption was diagnosed using clinical criteria^42^.

#### Psychiatric medication history

At each visit, participants underwent a detailed interview to assess prior medical history and medication use. Medications were then categorized according to whether there is a documented psychiatric indication for use.

### Statistical Approach

Prior to hypothesis testing, data were examined to evaluate missingness, identify outliers, and confirm that the data met analytic assumptions. Multivariate outliers in continuous variables were identified using Mahalanobis distance with a conservative chi-square criterion (*p*<.001, df=13). Descriptive statistics were computed to assess sample characteristics on variables of interest. All analyses were conducted in R Studio^43^ using R version 4.5.2.^44^

#### Identification of depressive symptom trajectories

Depressive symptom trajectory profiles were identified using latent class mixture modeling with class-specific time slopes using the ‘lcmm’ package in R.^45^ We estimated models with 2-5 classes to identify the best-fitting number of trajectory classes. Model selection was based on multiple criteria: (1) model fit indices, with lower values indicating better fit; (2) classification quality assessed by entropy (values >0.80 indicating strong class separation)^46^ and average posterior probabilities (values >0.7 indicating reasonable homogeneity within classes)^47^; (3) adequate size of smallest class (>5% of total sample); (4) model parsimony; and (5) clinical interpretability^47^. Once trajectory classes were identified, we compared participants across classes on sociodemographic and other characteristics. For continuous variables, we used one-way ANOVA for these comparisons, and for categorical variables used chi-square tests or Fisher’s exact tests in instances of small cell counts.

#### Relationship between depressive symptom trajectories and cardiovascular health

Once a latent class model was selected, we used structural equation modeling to assess the relationship between depressive symptom trajectories and CVH. Informed by prior research by Marsland and colleages^38^, CVH at V5 was modeled as a higher order latent factor indicated by intermediate latent factors: insulin resistance (glucose and insulin), adiposity (waist circumference and BMI), and dyslipidemia (triglycerides and non-HDL cholesterol), as well as observed systolic blood pressure (SBP; see supplemental methods and results for additional details). We adjusted for the following variables in the full structural model: length of time between the V3 and V5 assessment, maternal age, household income (dichotomized to >$60,000 or ≤$60,000, roughly the median household income level in 2010), education (dichotomized as having up to an associate degree vs. having a bachelor’s degree or higher), tobacco use history (dichotomized as yes or no), gravidity, APOs during the index pregnancy (dichotomized as present or absent), use of medications with a psychiatric indication during the index pregnancy or at V5 (dichotomized as yes or no), and pre-pregnancy BMI. These covariates were chosen given evidence of their impact on depressive symptoms, health behaviors and/or health outcomes in pregnancy and postpartum^48–52^. Maximum likelihood estimation with robust standard errors was used to accommodate categorical data (e.g., sociodemographic covariates)^53^ using the ‘lavaan’ package^54^ in R.

## Results

### Sample characteristics

The mean follow-up interval from delivery to the HHS visit (V5) was 3.2±0.9 years. Mean EPDS scores across timepoints were in the mild range (*M* range=5.5-5.7). At each timepoint, approximately 7% (range: 6.4-7.3%) of the sample scored at or above the clinical cut-off of 13. 15.5% exceeded this cutoff at one timepoint or more. See Supplemental Table 1 for additional self-reported demographic and descriptive information for the entire sample.

### Depressive symptom trajectory identification

Based on fit, class separation, class size, and interpretability, the four-class model was selected as the best-fitting model. Supplemental Table 2 includes model fit and classification statistics, and Supplemental Figure 1 depicts mean EPDS scores across time by class for each class model.

For the four-class model, the following EPDS patterns were identified: Group 1 (stable high symptom severity) was comprised of 212 (5.4%) of the sample and defined by having symptom scores ≥13 across all timepoints (range=13.8-14.1). Group 2 (decreasing symptom severity) included 476 participants (12.1%) who began pregnancy with elevated symptom scores (*M*=11.7±*SD*=2.7) but whose symptoms declined over time (V5 *M*=5.7±2.8). Group 3 (increasing symptom severity) was comprised of 848 participants (21.5%) who reported minimal symptoms early in pregnancy (*M=*6.4±2.4) but whose symptom scores worsened over time (V5 *M*=10.0±3.1). Group 4 (stable low symptom severity) was comprised of 2,411 (61.1%) participants who consistently reported minimal symptoms across time (range=3.3-3.5). These findings are presented in more detail in Table 1. EPDS scores over time for each of the four trajectory groups are presented in Figure 2.

**Figure 2.**
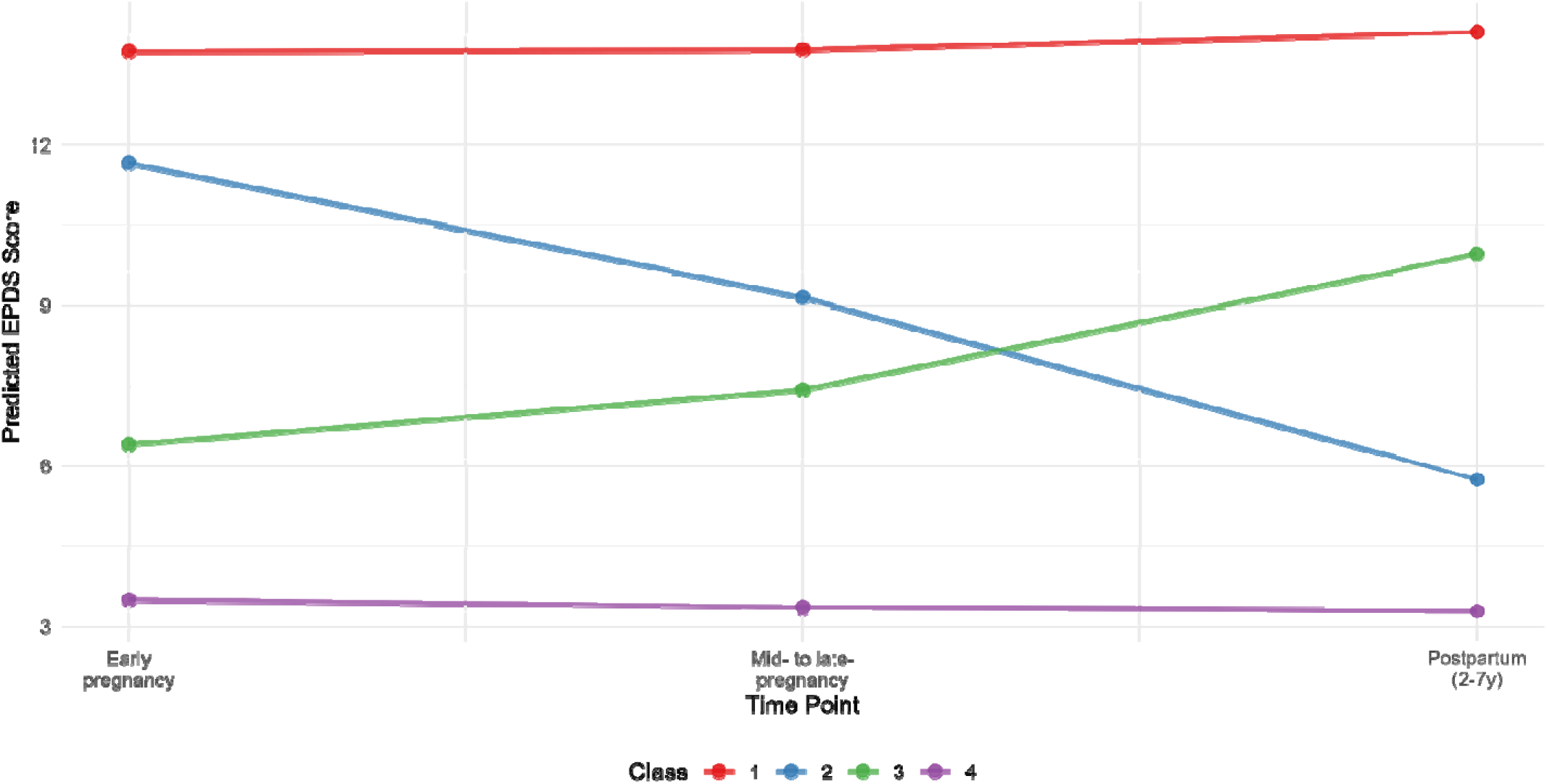
Predicted EPDS scores for each class across timepoints for the four-class model.

**Table 1.**
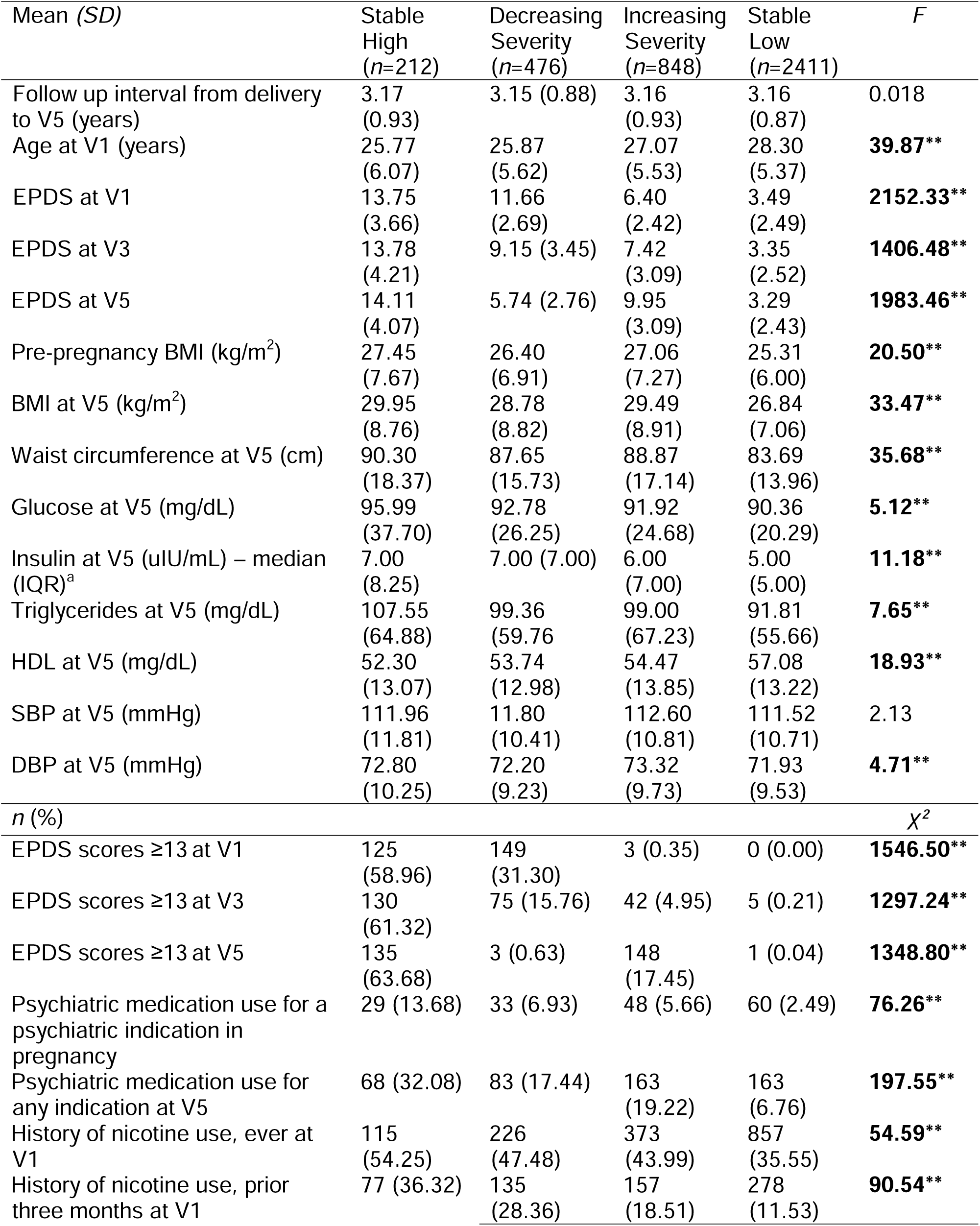
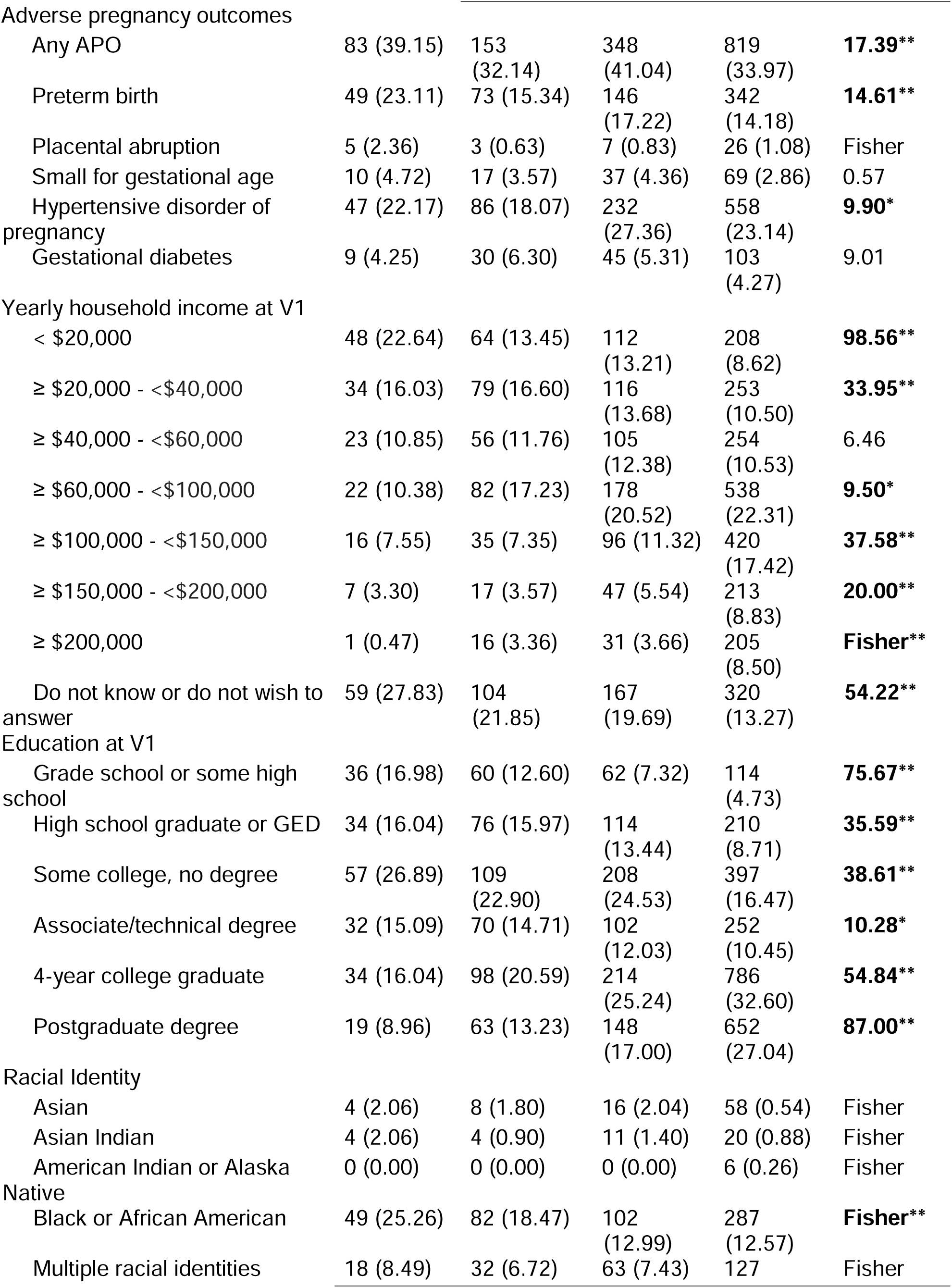
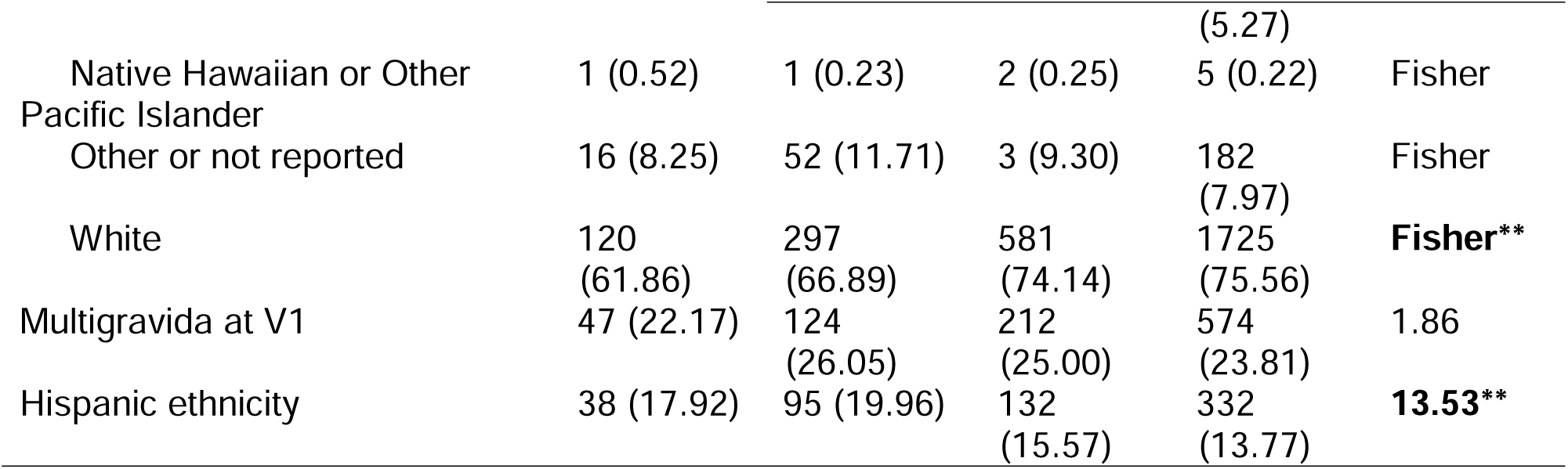
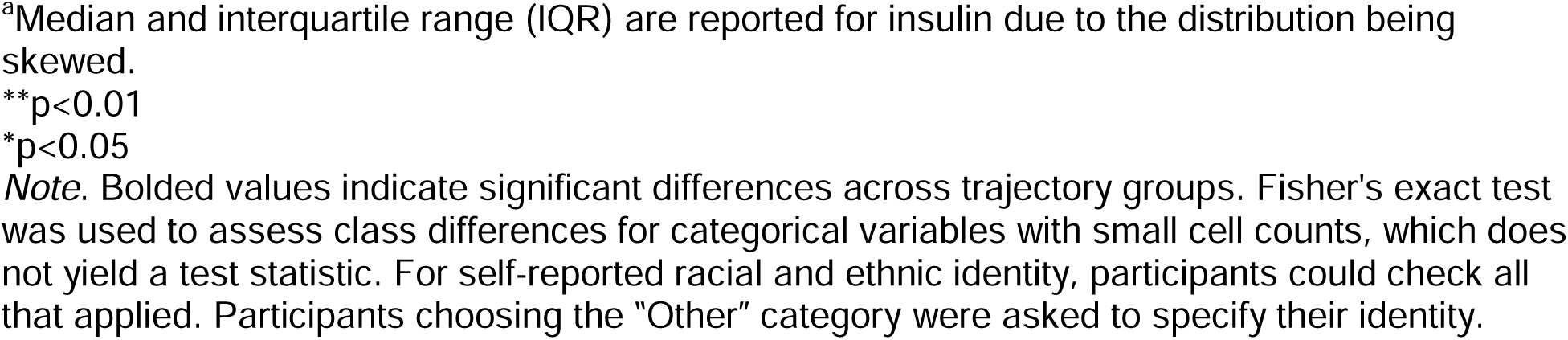
Demographic and clinical characteristics by EPDS class membership for the four-class.

### Full structural equation model

After confirming that the CVH measurement model adequately fit the data (see supplemental results for additional details), we conducted a full structural equation model with EPDS trajectory group membership predicting higher-order latent CVH at V5. The stable-low severity group was set as the comparison group.

The full structural model satisfactorily fit the data (χ² (88)=1254.7, p<0.01; robust RMSEA = 0.062, 90%CI=0.059 – 0.065; robust CFI=0.917; SRMR=0.054; R^2^ for CVH latent factor=0.19). Compared to the stable-low group, participants in the stable-high (β=0.06, *p*<0.01), decreasing (β=0.05, *p*=0.02), and increasing (β=0.08, *p*<0.01) severity groups all exhibited significantly worse CVH at V5, though effect sizes were small. Pairwise comparisons among the three elevated-symptom groups revealed no significant differences in latent CVH (all *p*s >0.24). Further, as documented in Table 1, values for most indicators included in the latent CVH construct were within normal limits across all groups, with the exception of body composition, which differed significantly across depressive symptom groups. Figure 3 displays the full structural model results. Model fit and effects did not change with the removal of outliers in sensitivity analyses (see supplemental methods and results for additional detail).

**Figure 3.**
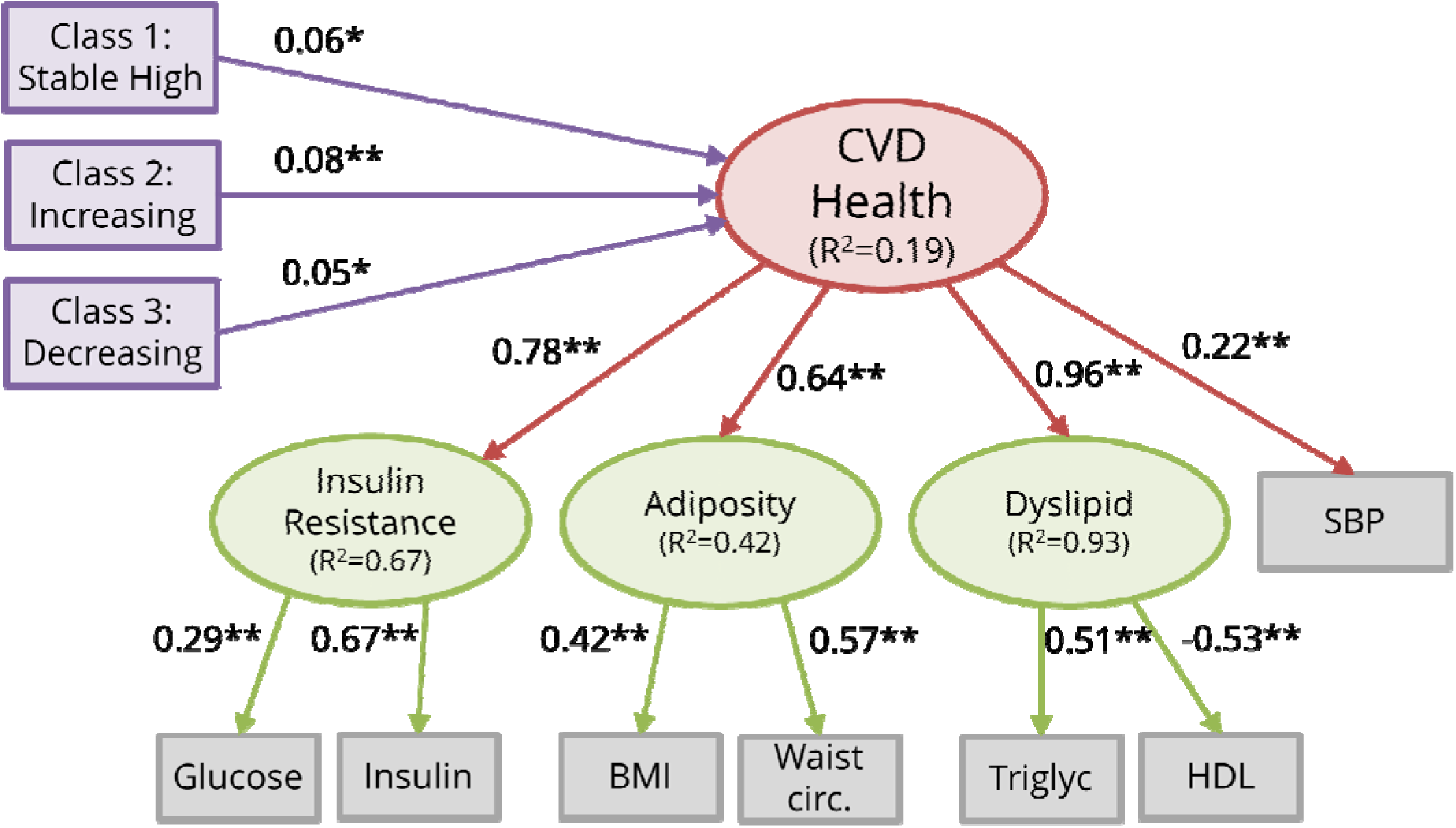
Depiction of the first structural model (χ² (88) = 1254.7, *p* = <0.01; robust RMSEA = 0.062, 90% confidence interval = 0.059 – 0.065; robust CFI = 0.917; SRMR = 0.054). Path coefficients are standardized. Interval between V3 and V5, age at V1, years of education at V1, household income at V1, tobacco use history reported at V1, gravidity, experience of APOs, use of a psychiatric medication during pregnancy, and use of a psychiatric medication at V5 were included as covariates for latent CVH. Additionally, pre-pregnancy BMI was regressed on V5 BMI and waist circumference. Note: \**p* <0.05, \*\**p* < 0.01. BMI = body mass index; CVD = cardiovascular disease; HDL = high density lipoprotein; SBP = systolic blood pressure.

## Discussion

Our findings demonstrate that even in a cohort where depressive symptom severity is low on average, those with consistently high, increasing, or initially high but then decreasing severity symptoms have worse CVH compared to individuals with consistently low severity symptoms. These findings extend prior research, which focused on individuals with a diagnosis of major depression^21,22,55^. While these previous studies provide critical evidence linking depression during pregnancy to future cardiovascular outcomes, their approaches did not consider individuals experiencing higher severity symptoms but who had not been diagnosed with major depression or individuals who had elevated symptoms below diagnostic thresholds.

Further, the current study extends previous findings by incorporating a third timepoint of depressive symptom data, measured 2-7 years post-delivery, and examining cardiovascular outcomes. In contrast, prior studies have typically modeled depression as a static exposure that does not account for the fact that there is significant inter-individual heterogeneity in the severity and course of depressive symptoms over time^26–28^ that may be relevant to identifying individuals at risk of CVD. One study that did evaluate symptom trajectories was performed in the same nuMoM2b cohort and demonstrated that a trajectory of worsening antenatal depressive symptoms was associated with risk of experiencing pre-term birth^10^, an APO that is associated with increased risk of future cardiovascular events^56^. Interestingly, in the present study, elevated symptom groups did not differ from one another with regard to CVH, suggesting that any elevation in symptoms during pregnancy and the extended postpartum period may contribute to poorer CVH. Finally, although prior studies demonstrated that depressive disorders in pregnancy are associated with increased risk of incident CVD, our findings indicate that the course of depressive symptoms from pregnancy through the extended postpartum period relate to continuous indicators of CVH prior to the onset of disease. Together, these results suggest there may be value in tracking depressive symptoms over time from pregnancy through the years post-delivery to identify individuals who may benefit from additional support for managing cardiovascular risk factors, even among those whose symptoms do not exceed established clinical thresholds.

It is important to note that the values for most CVH indicators in the current study were within normal limits for all depressive symptom trajectory groups, including among those with consistently elevated symptoms. The exceptions to this were the measures of body composition. At post-delivery follow-up, individuals in the stable-low group had a BMI of 27 kg/m^2^ while individuals in the increasing severity and stable-high symptom groups had a BMI near 30kg/m^2^, approximately a 10% difference. A similar pattern was identified for waist circumference. These findings are consistent with evidence that depression and obesity are bi-directionally related^57^, including during the perinatal period^50,58^, potentially due to shared underlying mechanisms such as systemic inflammation, autonomic dysregulation, and lower engagement in health behaviors^59^. Prior research has demonstrated that 5-10% reductions in weight are associated with improvements in cardiometabolic health^60,61^, and 5% weight loss is considered clinically significant^62^. Therefore, the magnitude of the differences in body composition observed across depressive symptom trajectory groups in the present study may translate to meaningful differences in future health risks. Further, the sample was relatively young at the time of the post-delivery assessment, around 31 years of age, well below the typical age of onset of CVD among women^63^. As such, it is possible that depression-associated risk profiles may worsen over time, particularly if individuals with higher severity symptoms do not receive appropriate treatment. Given that members of the nuMoM2b cohort continue to be followed and are now approaching midlife, it will be possible to evaluate this hypothesis when subsequent data are available.

### Strengths and limitations

The large size and prospective nature of the nuMoM2b-HHS cohort allowed us to evaluate and detect small but potentially meaningful relationships between depressive symptoms during pregnancy and the extended postpartum period and indicators of CVH. We were also able to use robust statistical approaches to identify depressive symptom subgroups who were heterogeneous with regard to both the severity and the course of their symptoms, providing an opportunity to assess how the course of symptoms over time related to cardiovascular outcomes. Nevertheless, there are also key limitations that should be taken into account when interpreting the results. First, there was considerable sample attrition between nuMoM2b and nuMoM2b-HHS, raising the possibility of selection bias that may limit the generalizability of findings. Relatedly, the sample reported higher education and income levels than the national average and may therefore have mental health and CVH profiles that differ from the general US population of perinatal individuals. Second, depressive symptoms were only measured three times over the study period and did not include assessment of symptoms during the immediate postpartum period, a time of significantly heightened vulnerability for onset or worsening of depression and associated comorbidities. Further, the final depressive symptom assessment was obtained concurrently with the CVH outcomes. As such, we were not able to address more nuanced questions about how the timing of depressive symptom changes relates to cardiovascular outcomes. Third, we did not account for subsequent pregnancies in our analyses, though there is evidence that cardiovascular risk accumulates across pregnancies^64^. Fourth, because the length of follow up in the nuMoM2b-HHS cohort was three years on average, we were unable to examine the relationship between depressive symptoms and longer-term cardiovascular endpoints such as myocardial infarction. Fifth, it is unclear whether the EPDS is the most appropriate measure of depressive symptoms beyond 12 months postpartum. Additional research is needed to determine the psychometric properties of the EPDS at longer post-delivery follow-up intervals. Finally, we were unable to account for pre-pregnancy depressive symptoms and therefore could not examine whether new onset of depression in pregnancy uniquely contributes to future cardiovascular risk, representing an important area for future research.

## Conclusions

Findings from this analysis of prospectively-collected data from a large multi-site cohort suggest that individuals with higher severity depressive symptoms at any point all exhibited worse CVH, regardless of the specific trajectory of symptoms. Additional research is critically needed to evaluate whether interventions targeting depression during pregnancy and the extended postpartum period have cardiovascular benefits, including delaying or preventing the onset of cardiovascular conditions later in life.

## Data Availability

Data is available by request via biodatacatalyst/numom2b.org

## Non-standard Abbreviations and Acronyms

APOs: adverse pregnancy outcomes
CFI: comparative fit index
CVD: cardiovascular disease
CVH: cardiovascular health
EPDS: Edinburgh Postnatal Depression Scale
RMSEA: root mean squared error of approximation
SRMR: standardized root mean squared residual

## Statement of Human Rights

All procedures performed were in accordance with the ethical standards of the institutional and/or national research committee and with the 1964 Helsinki declaration and its later amendments or comparable ethical standards.

## Independent Data Access and Analysis

Author SDD had full access to all of the data included in the present study, conducted all study analyses, and takes full responsibility for the integrity of the data and their analysis.

