## Supplement 1 for "Trajectories of depressive symptoms across pregnancy and the extended postpartum period and future cardiovascular health"

**SUMMARY**

This supplementary information includes more detailed descriptions of methods and results.

**Supplemental Methods**

**Measurement model estimation.** Consistent with previous research,^1^ cardiovascular health was modeled as a continuous higher order latent factor indicated by intermediate latent factors (i.e., insulin resistance, blood pressure, adiposity, and dyslipidemia). Fasting insulin and glucose served as observed indicators for the intermediate insulin resistance factor, systolic and diastolic blood pressure as observed indicators for the intermediate blood pressure factor, BMI and waist circumference as observed indicators of the intermediate adiposity factor, and HDL cholesterol and serum triglyceride levels as observed indicators of the intermediate dyslipidemia.

**Accounting for pre-pregnancy BMI in full structural model.** Modeling pre-pregnancy BMI as a covariate for latent cardiovascular health in full structural models produced a negative variance estimate for the adiposity intermediate latent factor, likely because it was highly correlated with other measures of body composition (r=0.81 for both V5 BMI and V5 waist circumference). Therefore, we regressed pre-pregnancy BMI directly onto V5 BMI and waist circumference rather than including as a covariate for higher-order latent cardiovascular health.

**Supplemental Results**

**Sample demographic characteristics.** eTable 1 includes the sociodemographic characteristics of the total sample.

**Comparison of lcmm models**. Indicators of model fit and classification were estimated for lcmm models with two to five classes. All fit statistics (AIC, BIC, and log-likelihood values) improved with each successive model. Classification quality was adequate across all models (entropy values all >0.7; posterior probability values all >0.8), indicating reasonable separation of classes for each model. However, the smallest class in the five-class model included only 105 participants, representing 2.7% of the sample, suggesting that this model may have over-fit the data. In addition, the five-class model had the lowest entropy (0.72) and posterior probability (0.8) values. eTable 2 includes model fit and classification estimates for the two, three, four, and five class models, and eFigure 2 displays EPDS scores across time for groups identified within each class model. Results favored a four-class solution.

**Cardiovascular health measurement model.** Prior to conducting full structural equation modeling, we first fit a measurement model to confirm that the proposed factor structure of cardiovascular health adequately fit the data. We initially specified a measurement model for cardiovascular health that included a higher order cardiovascular health latent factor indicated by four lower-order latent factors: insulin resistance (glucose and insulin), adiposity (BMI and waist circumference), dyslipidemia (triglycerides and HDL cholesterol), and blood pressure (systolic and diastolic blood pressure). However, the blood pressure latent factor produced a negative residual variance estimate for diastolic blood pressure, indicating a Heywood case. This estimation problem likely arose due to extreme multicollinearity between systolic and diastolic blood pressure (*r*=0.76) combined with their shared variance with other model components, leaving insufficient unique variance for diastolic blood pressure after accounting for the latent blood pressure factor.

To address this issue while retaining blood pressure in the model, we revised the measurement model to include only systolic blood pressure as a direct indicator of the higher-order cardiovascular health factor, alongside the intermediate insulin resistance, adiposity, and dyslipidemia latent factors. This approach resolved the estimation issue while maintaining the conceptual integrity of the cardiovascular health construct. The revised measurement model provided an adequate fit to the observed data (χ² (11)=161.40, *p*<0.01; robust RMSEA = 0.059, 90%CI=0.032 – 0.067; robust CFI=0.981; SRMR=0.026).

**Sensitivity analyses.** One hundred and sixty-four cases (4.2% of the sample) exceeded the threshold set for multivariate outlier detection using Mahalanobis distance. Therefore, we repeated analyses excluding these cases to evaluate the impact on findings. Latent class models produced a similar solution favoring the four-class model (AIC=61294.22; BIC=61369.08; log likelihood=-30635.11; mean posterior probability=0.85; entropy=0.75; smallest class size=206 (5.45%)), with trajectory groups in this model exhibiting similar EPDS patterns over time as was observed with the full sample. Full structural model fit indices were slightly attenuated but remained in the acceptable range (χ² (88)=1665.29, p<0.01; robust RMSEA = 0.071, 90%CI=0.068 – 0.074; robust CFI=0.914; SRMR=0.063). As was the case with the model using the full sample, the stable high (*β*=0.04, *p*=0.04), increasing severity (*β*=0.04, *p*=0.02), and decreasing severity (*β*=0.08, *p*<0.01) trajectory groups exhibited significantly lower cardiovascular health at V5 compared to the stable low trajectory group.

**eTable 1.** Demographic and clinical characteristics of sample.

|  | Mean (SD) |
| --- | --- |
| Follow up interval from delivery to V5 (years) | 3.16 (0.89) |
| Age at V1 (years) | 27.61 (5.56) |
| EPDS at V1 | 5.65 (4.18) |
| EPDS at V3 | 5.48 (4.15) |
| EPDS at V5 | 5.59 (4.32) |
| Pre-pregnancy BMI (kg/m^2^) | 25.93 (6.55) |
| BMI at V5 (kg/m^2^) | 27.81 (7.90) |
| Waist circumference at V5 (cm) | 85.63 (15.33) |
| Glucose at V5 (mg/dL) | 91.29 (23.06) |
| Insulin at V5 (uIU/mL) | 8.95 (11.16) |
| Triglycerides at V5 (mg/dL) | 95.10 (59.49) |
| HDL at V5 (mg/dL) | 55.86 (13.41) |
| SBP at V5 (mmHg) | 111.81 (10.77) |
| DBP at V5 (mmHg) | 72.30 (9.59) |
|  | n (%) |
| EPDS scores ≥13^a^ at V1 | 277 (7.0) |
| EPDS scores ≥13 at V3 | 252 (6.4) |
| EPDS scores ≥13 at V5 | 287 (7.3) |
| Psychiatric medication use for a psychiatric indication in pregnancy | 170 (4.31) |
| Psychiatric medication use for any indication at V5^b^ | 477 (12.09) |
| History of nicotine use, ever at V1 | 1571 (39.80) |
| History of nicotine use, prior three months at V1 | 647 (16.39) |
| Adverse pregnancy outcomes |  |
| Preterm birth | 610 (15.49) |
| Placental abruption | 41 (1.04) |
| Small for gestational age | 133 (3.37) |
| Hypertensive disorder of pregnancy | 923 (23.38) |
| Gestational diabetes | 187 (4.47) |
| Yearly household income at V1 |  |
| < $20,000 | 455 (11.53) |
| ≥ $20,000 - <$40,000 | 382 (9.68) |
| ≥ $40,000 - <$60,000 | 438 (11.10) |
| ≥ $60,000 - <$100,000 | 818 (20.72) |
| ≥ $100,000 - <$150,000 | 567 (14.37) |
| ≥ $150,000 - <$200,000 | 284 (7.19) |
| ≥ $200,000 | 253 (6.41) |
| Do not know or do not wish to answer | 650 (16.47) |
| Education at V1 |  |
| Grade school or some high school | 272 (6.89) |
| High school graduate or GED | 434 (10.99) |
| Some college, no degree | 771 (19.53) |
| Associate/technical degree | 456 (11.55) |
| 4-year college graduate | 1132 (26.68) |
| Postgraduate degree | 882 (24.35) |
| Racial Identity |  |
| Asian | 86 (2.32) |
| Asian Indian | 39 (1.05) |
| American Indian or Alaska Native | 6 (0.16) |
| Black or African American | 520 (14.03) |
| Multiple racial identities | 241 (6.11) |
| Native Hawaiian or Other Pacific Islander | 9 (0.24) |
| Other or not reported | 323 (8.72) |
| White | 2723 (73.48) |
| Multigravida at V1 | 957 (24.24) |
| Hispanic ethnicity | 597 (15.13) |

^a^A score of 13 or more on the EPDS is indicative of being at high-risk for a major depressive episode (Levis et al., 2020).

^b^Unlike the medical history interview administered at during pregnancy, the interview at V5 did not include questions regarding the reason for taking reported medications.

**eTable 2.** Fit and classification statistics for lcmm models with two to five classes

| Model | Log likelihood | AIC | BIC | Entropy | Mean posterior probability | Smallest class size, n (%) |
| --- | --- | --- | --- | --- | --- | --- |
| Two class | -32620.89 | 65253.77 | 65291.46 | 0.77 | 0.93 | 977 (24.75) |
| Three class | -32340.29 | 64698.58 | 64755.11 | 0.75 | 0.89 | 223 (5.65) |
| **Four class** | **-32219.56** | **64463.12** | **64538.49** | **0.74** | **0.85** | **212 (5.37)** |
| Five class | -32138.77 | 64307.54 | 64401.75 | 0.72 | 0.82 | 105 (2.66) |

*Note.* Bolded font denotes the identified best fitting model. For AIC and BIC model fit statistics, lower values indicate better fit. Entropy and mean posterior probability values ≥0.8 are considered indicative of good class separation, though values ≥0.7 are adequate. AIC=Akaike information criterion; BIC=Bayesian information criterion; lcmm=latent class mixture model.

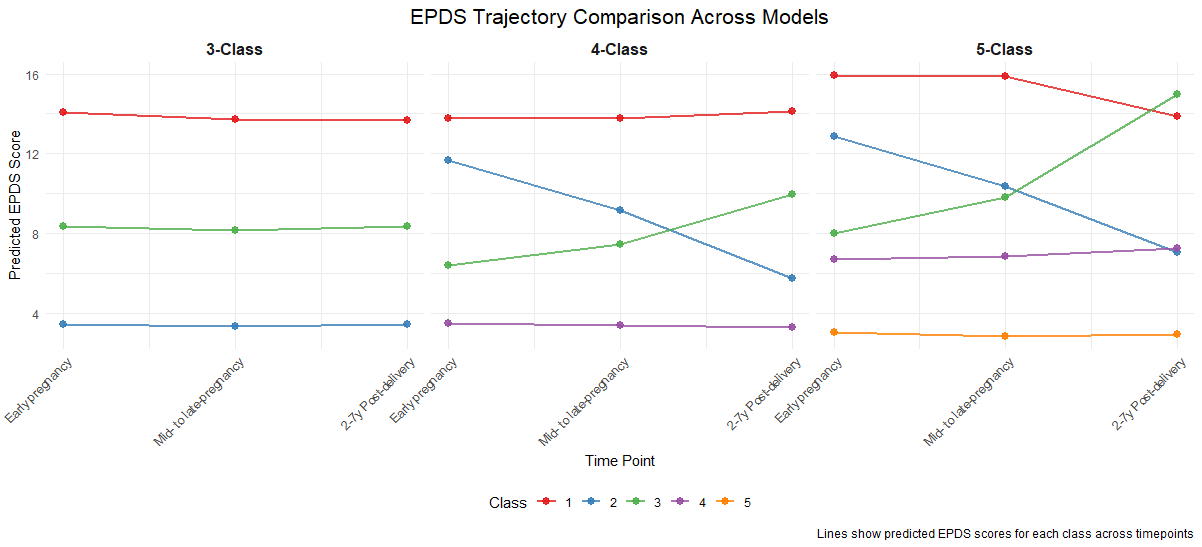

eFigure 1. Predicted EPDS scores for each class across timepoints for the three, four, and five class models.
